# Posttraumatic Stress Disorder Onset and Longitudinal Change in Biological Aging

**DOI:** 10.1101/2025.11.25.25340898

**Authors:** Kyle J. Bourassa, Melanie E. Garrett, Michelle Dennis, Annjanette Stone, Adrian C. Williams, VA Mid Atlantic MIRECC Workgroup, Jennifer C. Naylor, Allison E. Ashley-Koch, Nathan A. Kimbrel, Jean C. Beckham

**Author notes:** Please direct correspondence to Dr. Kyle J. Bourassa, 508 Fulton Street, Durham, NC 27705. **Group information:** The VA Mid-Atlantic MIRECC Workgroup contributors include: Patrick S. Calhoun, PhD, Eric Dedert, PhD, Eric B. Elbogen, PhD, Robin A. Hurley, MD, Jason D. Kilts, PhD, Angela Kirby, MS, Scott D. McDonald, PhD, Sarah L. Martindale, Ph.D, Christine E. Marx, MD, MS, Scott D. Moore, MD, PhD, Rajendra A. Morey, MD, MS, Jared A. Rowland, PhD, Robert D. Shura, PsyD, Cindy Swinkels, PhD, H. Ryan Wagner, PhD. **Data Sharing Statement:** Data from the Post Deployment Mental Health (PDMH) Study are available to researchers who request access through the VISN 6 MIRECC and follow the appropriate data access protocols.

## Abstract

People with posttraumatic stress disorder (PTSD) are at increased risk for poor health, which could be explained by faster rates of biological aging. However, associations between PTSD and aging have most often been investigated using cross-sectional designs, with few longitudinal studies using more recently developed epigenetic measures of aging. To test whether changes in PTSD status were associated with changes in biological aging over roughly 12 years, we used data from 400 veterans assessed at two visits in the Post-Deployment Mental Health Study. Biological aging was assessed by DunedinPACE, with additional results shown for PC-GrimAge and PC-PhenoAge. Between two occasions spanning an average of 11.9 years, veterans who developed new onset PTSD showed significant increases in DunedinPACE (β = 0.24, 95% CI [0.07, 0.41], *p* = .006), whereas remission in PTSD between occasions was not associated with significant decreases in the rate of aging (β = −0.13, 95% CI [-0.33, 0.08], *p* = .207). When assessing PTSD symptoms, increases in PTSD symptoms between baseline and follow-up were associated with increases in DunedinPACE over the same period (β = 0.07, 95% CI [0.01, 0.14], *p* = .032), and vice versa. Estimates for PC-GrimAge largely replicated those for DunedinPACE, whereas PC-PhenoAge replicated some associations.

**Conclusions:** These results suggest that changes in PTSD are associated with longitudinal changes in biological aging. Efforts to prevent the onset of PTSD and reduce PTSD symptoms could slow aging and reduce risk for poor health.

## Introduction

Posttraumatic stress disorder (PTSD) is associated with increased risk for a wide range of chronic diseases, including cardiovascular^1–3^, pulmonary^4^, and metabolic diseases^5,6^. This increased risk extends to other clinical outcomes, including premature death^1,7^. Recent studies have suggested that accelerated biological aging could help explain health consequences associated with PTSD^8,9^, as individuals with PTSD are generally aging more quickly than people without PTSD^10^ and faster aging is associated with greater risk for chronic disease and mortality^11^. Intervening to treat PTSD or the health-relevant sequelae of PTSD might slow aging and reduce risk for poor health among people who experience trauma and develop PTSD^9^, however, this possibility requires additional support from longitudinal studies that can show that changes in PTSD are associated with changes in aging over time. This kind of empirical evidence could support future randomized controlled trials (RCTs) aiming to provide causal evidence as to which treatments might reduce the health consequences of PTSD.

Although much of the evidence linking PTSD to accelerated aging is cross-sectional, some recent studies have investigated longitudinal changes in PTSD and aging with mixed results^12–14^. For example, a recent meta-analysis of 7 cohorts found an association between new onset PTSD and a first-generation epigenetic clock trained on chronological age (i.e., Horvath), but no association with a second-generation measure of aging trained on mortality (i.e., GrimAge)^12^. Another recent longitudinal study did not find evidence that changes in PTSD status were associated with changes in six epigenetic measures of aging^13^. Given the mixed current evidence linking PTSD and change in biological aging, there is a need for additional studies that can examine associations between PTSD and biological aging over long follow-ups using gold standard clinical interviews to diagnose PTSD. Similarly, given the variety of epigenetic measure of aging that have been developed, it would also be valuable if new studies could use epigenetic measures that were developed to be more sensitive to changes in aging. This includes third-generation measures trained on longitudinal change in biomarkers (DunedinPACE^15^) in order to index biological *aging* rather than biological *age*. Similarly, second-generation aging measures developed using principle components to increase reliability (i.e., PC-GrimAge PC-PhenoAge) may better able to detect change when assessing aging over multiple occasions^16^. To do so, we examined whether changes in PTSD status and PTSD symptoms were associated with changes in biological aging over 11.9 years for 400 post-9/11 veterans^17^.

## Methods and Materials

### Participants and Study Design

Participants included 400 veterans enrolled in the VISN 6 Mid-Atlantic Mental Illness Research, Education, and Clinical Center’s (MIRECC’s) Post Deployment Mental Health (PDMH) Study^17^ with DNAm data generated at the baseline and follow-up (*N* = 400; e**Figure 1** provides a flowchart of inclusion and exclusion criteria, **eTable 1** provides comparisons of demographic and PTSD characteristics for the sample). The Durham, Richmond, W.G. Bill Hefner VA and Central Virginia VA Health Care Systems’ Institutional Review Boards approved the study protocol and participants provided informed consent.

**Figure 1.**
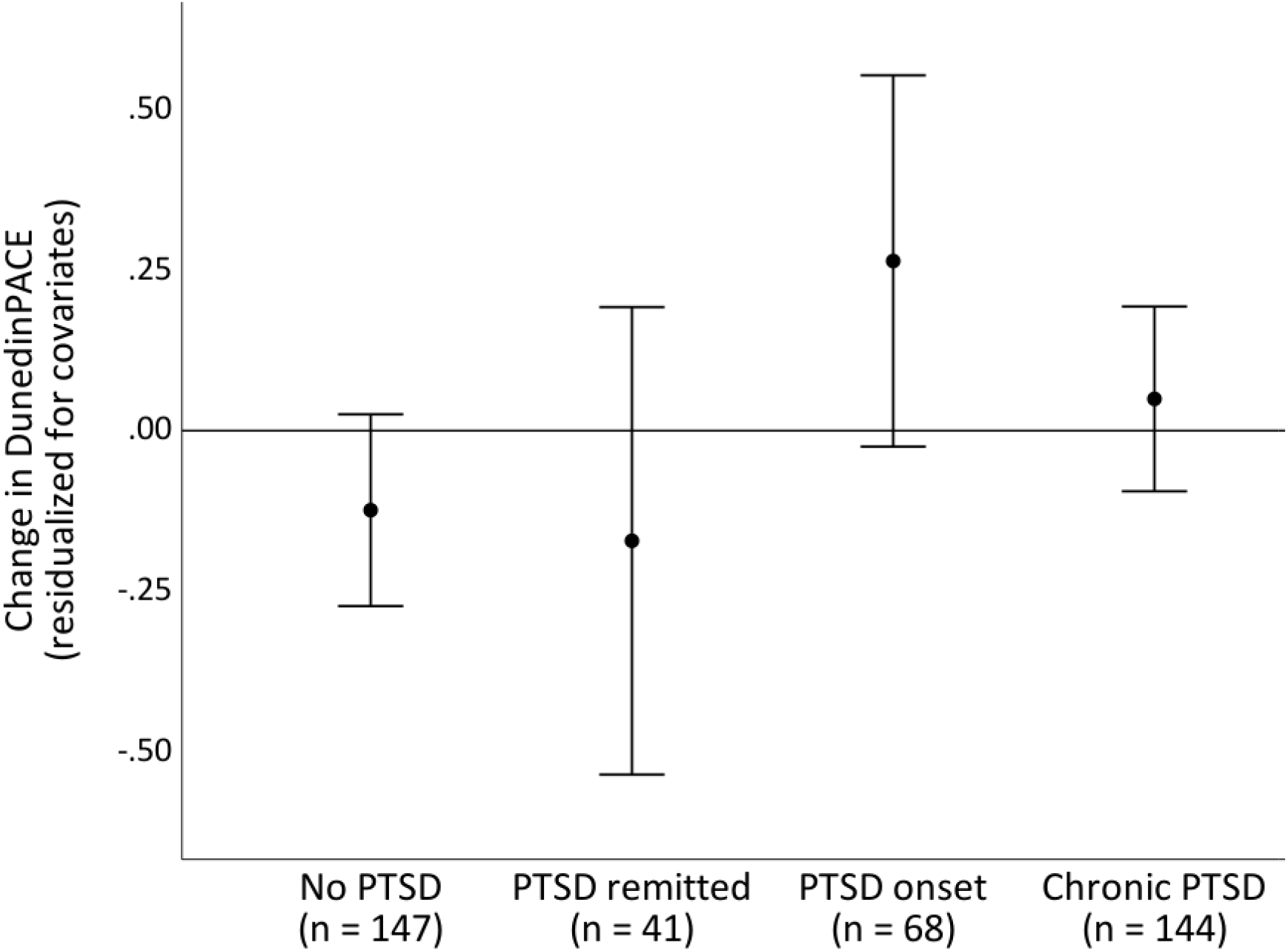
Visualization of change in DunedinPACE among the four groups representing change in PTSD diagnosis. Veterans were categorized based on longitudinal PTSD status: 1.) no PTSD at baseline or follow-up = No PTSD; 2.) PTSD at baseline but not follow-up = PTSD remitted; 3.) No PTSD at baseline but PTSD at follow-up = PTSD onset; 4.) PTSD at both occasions = Chronic PTSD. DunedinPACE aging scores were residualized for demographic and technical covariates, as well as baseline DunedinPACE to represent residualized change in aging scores. Errors bars represent 95% confidence intervals.

### Measures

#### Biological aging

Whole blood was collected during PDMH baseline and follow up assessments and analyzed using the Infinium MethylationEPIC v1.0 Beadchip at baseline and MethylationEPIC Beadchip v2.0 at follow up (Illumina Inc., San Diego, CA). Sample quality control was performed using the minfi^12^ R package. Samples were excluded if average fluorescence signal intensity was below 2000 arbitrary units or <50% of the mean intensity of all samples, >10% of probes were not detectable (p-value > 0.001), if a sex mismatch was detected, or if the sample was deemed an outlier on principal component analysis plots. One follow-up sample had a 96.8% probe failure rate and their aging values were coded as missing. Probe quality control and data normalization were performed within each batch using the R package SeSAMe^18^. Specifically, probes with poor design were masked and color channels were inferred (for Infinium I probes) followed by dye bias correction and noob background subtraction. Detection p-values were generated using the pOOBAH method; probes not detected (detection p-value > 0.001) in >10% of samples and those hybridizing to multiple locations in the genome were removed. Beta values for duplicated EPIC v2.0 probes were collapsed to the mean to allow for comparison with EPICv1 probes. Principal component analysis was used to assess clustering of values by experimental batch and adjustments were completed using ComBat in the R package sva^19^. Methylation values reflected the resulting normalized and adjusted beta values derived from methylated DNA. As noted elsewhere, EPIC v1.0 and 2.0 chips produce highly correlated results^20^. PC clocks were residualized on chronological age. DunedinPACE, PC-GrimAge, and PC-PhenoAge scores were generated at both occasions using published algorithms^15,16^. Baseline and follow-up DunedinPACE (*r* = .76, *p* < .001) and PC-GrimAge (*r* = .78, *p* < .001) aging scores were highly correlated, whereas PC-PhenoAge were moderately corrected (*r* = .51, *p* < .001).

#### White blood cell type proportions

The FlowSorted.Blood.EPIC R package was used to estimate white blood cell type proportions (T lymphocytes [CD4+ and CD8+], B cells [CD19+], monocytes [CD14+], NK cells [CD56+]) using publicly available reference datasets^21^.

#### Posttraumatic stress disorder (PTSD)

PTSD was assessed at baseline and follow-up using the Structured Clinical Interview based on *DSM-IV* criteria^22^ and self-reported PTSD symptoms were assessed using the Davidson Trauma Scale^23^ (DTS). Veterans were assigned to current PTSD at each occasion if they met interviewer-rated criteria for PTSD or had a DTS score above the clinical cutoff of 35^10,24^. DTS scores were also used to assess PTSD symptoms at both occasions. Dummy codes were created to represent PTSD onset (coded 0 = not present, 1 = present) and PTSD remission (coded 0 = not present, 1 = present) for models assessing change in PTSD status.

#### Demographics

Participants reported their age, sex, race, ethnicity, and years of education. Sex, race, and ethnicity were confirmed by genetic data^10^. Time between assessments was also calculated and included as a covariate in all models.

#### Smoking

Veterans reported their smoking status (0 = never smoker, 1 = past smoker, 2 = current smoker) at follow-up, which were confirmed using reported smoking at baseline.

### Data Analysis

We used multiple linear regression models to examine whether changes in PTSD status (PTSD onset; PTSD remission coded) were associated with change in DunedinPACE scores from baseline to follow-up. We also modeled whether change in PTSD symptoms from baseline to follow-up was associated with change in DunedinPACE, to test whether faster aging might be a risk factor to develop future PTSD. All models included a number of covariates, including demographics (age at follow-up, sex, race and ethnicity, years of education, and time between study occasions), technical covariates (estimated white blood cell count proportions at baseline and follow-up), smoking, and baseline measures of PTSD (PTSD diagnosis or PTSD symptoms at baseline). Models also included baseline measures of DunedinPACE to produce a residualized regression model of change in DunedinPACE. We also tested whether baseline aging scores predicted change in PTSD and show associations for PC-GrimAge and PC-PhenoAge. Multiple regression models were run in MPLUS v8.3^25^ using full maximum likelihood to account for missing data. β estimates represent standardized effects.

## Results

The sample included 400 veterans (84.8% men) who were 39.3 years old at baseline and 51.2 years old at follow-up, averaging 11.9 years between study visits. 202 (50.5%) veterans were non-Hispanic Black, whereas 198 (49.5%) were non-Hispanic White. Demographics are presented in **eTable 2**.

### Longitudinal PTSD and DunedinPACE Associations

Veterans who developed PTSD between study occasions showed significant increases in DunedinPACE the 11.9 years between study occasions (β = 0.24, 95% CI [0.07, 0.41], *p* = .006), however, PTSD remission was not associated with slowed aging over the same period (β = −0.13, 95% CI [-0.33, 0.08], *p* = .207). Changes in PTSD symptoms were associated with changes in DunedinPACE between baseline and follow-up (β = 0.07, 95% CI [0.01, 0.14], *p* = .032; **Table 1**), such that increases in PTSD symptoms were associated with increases in DunedinPACE and vice versa.

**Table 1.** Association of PTSD and epigenetic measures of aging.

| *N* = 400 | DunedinPACE at follow-up |  | PC-GrimAge at follow-up |  | PC-PhenoAge at follow-up |  |
| --- | --- | --- | --- | --- | --- | --- |
| | $\beta$ | 95% CI | $\beta$ | 95% CI | $\beta$ | 95% CI |
| **PTSD diagnostic status** |  |  |  |  |  |  |
| PTSD onset | 0.24** | [0.07, 0.41] | 0.15* | [0.01, 0.28] | 0.26* | [0.05, 0.46] |
| PTSD remission | -0.13 | [-0.33, 0.07] | -0.13 | [-.029, 0.04] | -0.03 | [-0.21, 0.28] |
| Baseline PTSD status | 0.08 | [-0.06, 0.22] | 0.12* | [0.01, 0.23] | 0.16 | [-0.01, 0.32] |
| Baseline aging score | 0.76** | [0.71, 0.80] | 0.70** | [0.65, 0.75] | 0.58** | [0.53, 0.64] |
| Age | -0.05 | [-0.12, 0.02] | -0.01 | [-0.06, 0.03] | -0.07* | [-0.14, -0.01] |
| Sex | -0.09 | [-0.25, 0.08] | -0.26** | [-0.40, -0.12] | 0.03 | [-0.17, 0.23] |
| Race and ethnicity | -0.35** | [-0.49, -0.21] | 0.05 | [-0.07, 0.16] | -0.40** | [-0.57, -0.23] |
| Years of education | 0.02 | [-0.03, 0.08] | -0.00 | [-0.04, 0.04] | -0.01 | [-0.07, 0.05] |
| Smoking | 0.05 | [-0.01, 0.10] | 0.03 | [-0.02, 0.08] | -0.04 | [-0.10, 0.02] |
| Years between occasions | 0.06 | [-0.00, 0.11] | 0.06** | [0.02, 0.10] | 0.04 | [-0.03, 0.09] |
| **PTSD symptoms** |  |  |  |  |  |  |
| PTSD symptom change | 0.07* | [0.01, 0.14] | 0.07** | [0.02, 0.19] | 0.05 | [-0.01, 0.12] |
| Baseline PTSD symptoms | 0.03 | [-0.04, 0.09] | 0.06* | [0.01, 0.12] | 0.05 | [-0.01, 0.12] |
| Baseline aging score | 0.77** | [0.72, 0.82] | 0.70** | [0.65, 0.75] | 0.58** | [0.52, 0.63] |
| Age | -0.05 | [-0.12, 0.02] | -0.01 | [-0.06, 0.03] | -0.07* | [-0.14, -0.01] |
| Sex | -0.10 | [-0.26, 0.07] | -0.26** | [-0.39, -0.12] | 0.02 | [-0.18, 0.22] |
| Race and ethnicity | -0.36** | [-0.50, -0.22] | 0.05 | [-0.06, 0.17] | -0.41** | [-0.58, -0.24] |
| Years of education | 0.02 | [-0.04, 0.08] | -0.00 | [-0.05, 0.04] | -0.01 | [-0.07, 0.05] |
| Smoking | 0.05 | [-0.01, 0.11] | 0.03 | [-0.02, 0.08] | -0.04 | [-0.09, 0.03] |
| Years between occasions | 0.06 | [-0.00, 0.12] | 0.06** | [0.02, 0.10] | 0.03 | [-0.03, 0.09] |

*Note:* All models included demographic (sex, race and ethnicity, education, and time between study occasions), technical covariates (white blood cell proportions), baseline aging measures and technical covariates, as well as baseline measures of PTSD (PTSD diagnostic status or PTSD symptoms). Baseline and follow-up white blood cell proportions were not included for readability. All $\beta$ s reported reflect standardized effect sizes. CI = confidence interval.
\* $p < .05$ . \*\* $p < .01$ .

We next assessed whether initial DunedinPACE levels might predict future onset of PTSD. Baseline DunedinPACE was not associated with future PTSD diagnostic status (β = −0.02, 95% CI [-0.12, 0.08], *p* = .700) or change in PTSD symptoms (β = −0.08, 95% CI [-0.18, 0.01], *p* = .094).

### Secondary Analyses: Results for PC-GrimAge and PC-PhenoAge

Estimates for PC-GrimAge largely replicated those for DunedinPACE with similar effect sizes for changes in PTSD status and symptoms. PTSD onset was also associated with increases in PC-PhenoAge, whereas change in PTSD symptoms was not (**Table 1**).

## Discussion

In this longitudinal study, PTSD onset was associated with increases in biological aging over an 11.9-year follow-up. The patterns of initial levels and change in aging observed among veterans with different PTSD statuses reflected these findings (**Figure 2**). Veterans without PTSD had the slowest aging and veterans with chronic PTSD (i.e., PTSD at both baseline and follow-up) had faster aging at baseline and follow-up. We did not, however, find evidence that veterans with faster aging were more likely to develop PTSD^7^, suggesting that aging reflects changes in physiology after trauma, rather than predisposing individuals to develop PTSD. Although PTSD remission was not associated with slower aging, the results highlight the importance of testing the association between change in PTSD status and biological aging in larger samples with more statistical power.

**Figure 2.**
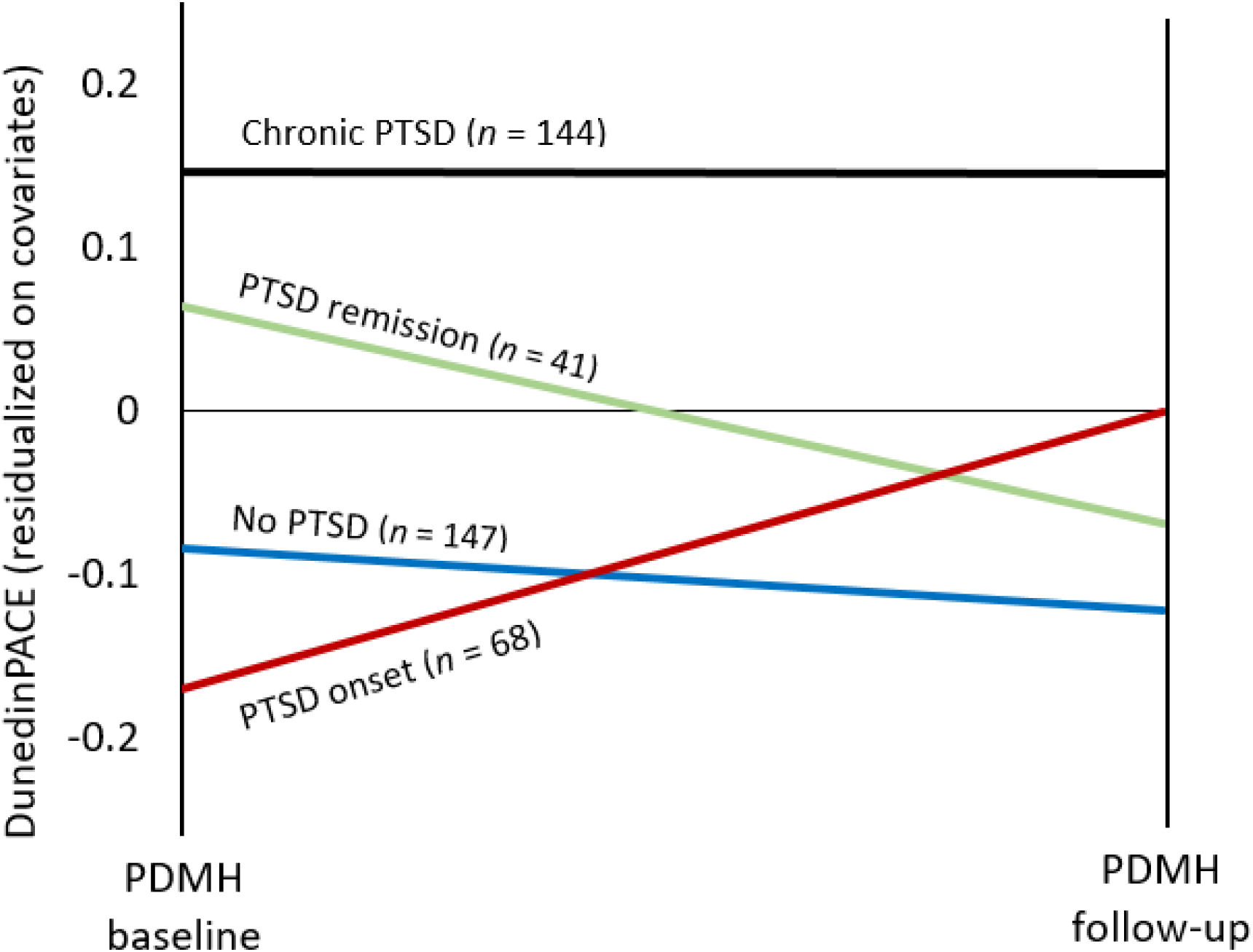
Initial levels and change in DunedinPACE scores among the PTSD groups. Figure 1 presents the results with associated confidence intervals, which were excluded from this figure to maintain legibility.

Our results have several implications. First, these findings provide evidence that PTSD is associated with second- and third-generation epigenetic measures of aging, and extend these results to longitudinal models. These results stand in contrast to other studies that did not find such associations^13^, which suggests the need for additional studies to replicate our results using larger samples and varying populations (e.g., military and non-military). Our study is notable for the relatively long period (11.9 years, on average) between assessments of PTSD and DunedinPACE. Second, our results highlight the potential value in preventing the onset of PTSD and using behavioral or pharmacological treatments to reduce symptoms of PTSD. If these treatments can slow biological aging, they might also reduce the risk for chronic disease and premature mortality associated with PTSD^1,2,5^. Future studies would benefit from linking PTSD, aging, and health in larger cohorts to provide further evidence that longitudinal change in aging scores can be assessed reliably. Ideally, RCTs with long-term follow up would also be used to determine the extent to which PTSD treatment might reduce risk for poor health.

Our study included several limitations. First, estimates were derived from a longitudinal subsample of the PDMH^11^ and will require replication in the full cohort once DNAm data is generated in more veterans, as well as replication in other cohorts. Second, our DNAm data was from two versions of methylation chips that reflected changes in manufacturing, which meant chip version was confounded with baseline and follow up sampling. Though we note that the PC clocks are generally more reliable across chips types^16^ and DunedinPACE is highly consistent between EPIC v1.0 and v2.0 chips^20^, this change in chip could have affected our results. Third, our study used DSM-IV criteria for PTSD diagnosis. Although these were the criteria when the baseline PTSD assessment was conducted, it is possible these results might be differ if using DSM-5 criteria. Finally, it is unclear to what extent the results from post-9/11 veterans might generalize to veterans from other eras of service or non-veteran populations.

## Conclusions

We found PTSD onset and change in PTSD symptoms were associated with increases in the rate of biological aging over 12 years in a sample of 400 U.S. military veterans. These results suggest that efforts to prevent the onset of PSTD or treat PTSD symptoms have the potential to slow aging and improve health among individuals who experience traumatic events and develop PTSD.

## Data Availability

Data from the Post Deployment Mental Health (PDMH) Study are available to researchers who request access through the VISN 6 MIRECC and follow the appropriate data access protocols.

