## Supplement for "Posttraumatic Stress Disorder Onset and Longitudinal Change in Biological Aging"

**Supplemental Materials**

**eFigure 1.** ….…….…….…………….………………………….……...……………………………………………..…...…….………….……...2

**eTable 1.** ….…….…….…………….………….…….….…………………………………………………………….………………….....……...3

**eMethod 1.** ….…….…….…………………………………….……...……………………………………………..…...…….………….……...4

**eReferences 1.**…….…….…………………………………….……...……………………………………………..…...…….………….……...5

**eTable 2.** ….…….…….…………….………….…….….…………………………………………………………….………………….....……...6

PDMH Repository

N = 3,876

Exclusions:

DNAm not assayed at baseline: N = 1,567

Baseline DNAm sample

N = 2,309

Exclusions:

DNAm not assayed at follow-up: N = 1,909

Analysis Cohort

N = 400

***eFigure 1.*** CONSORT-style diagram showing the selection of the final analysis cohort from the original PDMH repository for the current study. The number of veterans selected for the DNAm subsample at baseline was based on the availability of grant funds for DNA methylation processing. Selection of veterans for the DNAm follow-up sample required having an available blood sample and the Illumina EPIC V1 chip at baseline, resulting in the final analysis cohort.

**eTable 1.** Comparing the selected sample to participants with DNAm data at baseline but not follow-up.

|  | **Analytic sample**  ***(n* = 400)** | | **Excluded veterans with DNAm *(n* = 1,909)** | |  |
| --- | --- | --- | --- | --- | --- |
|  | ***Mean / %*** | ***SD*** | ***Mean / %*** | ***SD*** | ***Difference between samples (Cohen’s d)*** |
| Baseline Age | 39.3 | 9.9 | 37.0 | 10.1 | 0.23*** |
| Sex | 84.8% |  | 77.5% |  | -0.18*** |
| Race and ethnicity | 50.5% |  | 47.5% |  | -0.06 |
| Baseline education | 13.6 | 3.5 | 13.6 | 3.6 | 0.01 |
| Current smoking at baseline | 57.7% |  | 50.7% |  | 0.14* |
| Current PTSD at baseline | 46.3% |  | 51.5% |  | -0.11 |
| Baseline PTSD symptoms | 40.3 | 38.3 | 43.5 | 40.2 | -0.08 |
| *Note:* Sex reported (%) men, race and ethnicity reported (%) non-Hispanic White, with remaining percent non-Hispanic Black. If data was missing, means and percentages were calculated using observed values.  ** = p* < .05; ** = *p <* .01; *** = *p <* .001 | | | | | |

**eMethod 1. DNA methylation data quality control and normalization.**

Whole blood was collected during PDMH baseline and follow up assessments and analyzed using the Infinium MethylationEPIC v.1.0 Beadchip at baseline (Illumina Inc., San Diego, CA) and MethylationEPIC Beadchip v2.0 at follow up. Sample quality control was performed using the minfi^1^ R package. Samples were excluded if average fluorescence signal intensity was below 2000 arbitrary units or <50% of the mean intensity of all samples, >10% of probes were not detectable (p-value > 0.001), if a sex mismatch was detected, or if the sample was deemed an outlier on principal component analysis plots. In total, 1 sample was removed due to quality control (96.8% probe failure rate) and included with their values coded as missing. Cell type proportions were estimated using the FlowSorted.Blood.EPIC R package with publicly available reference datasets^2^. Probe quality control and data normalization were performed within each batch using the R package SeSAMe^3^. Specifically, probes with poor design were masked and color channels were inferred (for Infinium I probes) followed by dye bias correction and noob background subtraction. Detection p-values were generated using the pOOBAH method; probes not detected (detection p-value > 0.001) in >10% of samples and those hybridizing to multiple locations in the genome were removed. Beta values for duplicated EPICv2 probes were collapsed to the mean to allow for comparison with EPICv1 probes. Principal component analysis was used to assess clustering of values by experimental batch and adjustments were completed using ComBat in the R package sva^4^. Methylation values reflected the resulting normalized and adjusted beta values derived from methylated DNA. As noted elsewhere, EPIC v1.0 and 2.0 chips produce highly correlated results^5^. PC clocks were residualized on chronological age.

**eReferences 1.**

**eTable 2.** Full demographic characteristics of the sample

| ***N* = 400** | ***Mean (SD)*** | ***%*** |
| --- | --- | --- |
| Baseline age | 39.3 (9.9) |  |
| Follow-up age | 51.2 (9.9) |  |
| Time between occasions | 11.9 (0.6) |  |
| Sex |  | 84.8% men |
|  |  | 15.3% women |
| Race and ethnicity |  | 50.5% non-Hispanic Black |
|  |  | 49.5% non-Hispanic White |
| Years of education | 13.6 (3.5) |  |
| Smoking |  | 57.5% never smoker |
|  |  | 20.0% past smoker |
|  |  | 22.5% current smoker |
| Baseline PTSD |  | 46.3% PTSD diagnosis |
| Follow-up PTSD |  | 47.0% PTSD diagnosis |
| Baseline PTSD symptoms | 40.3 (38.3) |  |
| Follow-up PTSD symptoms | 42.3 (37.7) |  |
| *Note:* PTSD = posttraumatic stress disorder. | | |
